# Influence of socioeconomic status on the presence of obstructive coronary artery disease and cardiovascular outcomes in patients undergoing invasive coronary angiography

**DOI:** 10.1101/2023.03.08.23287013

**Authors:** Jaehoon Chung, Hack-Lyoung Kim, Hyun Sung Joh, Woo-Hyun Lim, Jae-Bin Seo, Sang-Hyun Kim, Joo-Hee Zo, Myung-A Kim

**Affiliations:** Division of Cardiology, Department of Internal Medicine, National Medical Center, Seoul, Republic of Korea; Division of Cardiology, Department of Internal Medicine, Boramae Medical Center, Seoul National University College of Medicine, Seoul, Republic of Korea

**Keywords:** cardiovascular disease, coronary angiography, coronary artery disease, medical aid beneficiary, national health insurance beneficiary, socioeconomic status

## Abstract

**Background:** Although it has been documented that low socioeconomic status (SES) is associated with increased risk of mortality and cardiovascular disease (CVD), this issue has not been addressed in patients at high CVD risk. This study was performed to investigate the association of the patient’s SES with the presence of obstructive coronary artery disease (CAD) and long-term cardiovascular outcomes who undergo invasive coronary angiography (ICA).

**Methods:** A total of 9,530 patients who underwent ICA for the evaluation and treatment of CAD (66.0±12.3 years and 60.2% male) were retrospectively reviewed. The patients were divided into two groups according to the health insurance type: those with low SES who had the Medical aid program (Medical aid beneficiary [MAB] group; n=1,436) and those with high SES who had the National Health Insurance program (National Health Insurance beneficiary [NHIB] group; n=8,094). The primary outcome was a composite of cardiac death, acute myocardial infarction, coronary revascularization, and ischemic stroke.

**Results:** Of the study patients, 1,436 (15.1%) were in the MAB group. The prevalence of cardiovascular risk factors was higher in the MAB group compared to the NHIB group. However, the prevalence of obstructive CAD was similar between the two groups (62.8% vs. 64.2%; *P*=0.306). During a median follow-up period of 3.5 years (interquartile range, 1.0 to 5.9 years), the incidence of the composite cardiovascular event was significantly higher in the MAB group than in the NHIB group (20.2% vs. 16.2%, *P*<0.001). In multivariable Cox regression analysis, compared to the NHIB group, the MAB group was independently associated with worse clinical outcomes even after controlling for potential confounders (adjusted odds ratio, 1.28; 95% confidence interval, 1.07 to 1.54; *P*=0.006).

**Conclusions:** Although CAD prevalence was similar, MABs showed an increased risk of composite cardiovascular events than NHIBs in Korean adults undergoing ICA. This provides additional evidence for the association between low SES and an increased risk of CVD, even in high-risk subjects.

**Clinical perspective:** *What is new?:*

- This study provided evidence for a relationship between low socioeconomic status and increased cardiovascular disease risk in a high-risk population.

*What are the clinical implications?:*

- Subjects with low socioeconomic status are more likely to develop cardiovascular disease and to have a higher frequency of related risk factors, so cardiovascular disease tests and treatments should be performed more aggressively.
- Appropriate risk stratification of low socioeconomic status patients with traditional risk factors for cardiovascular disease is important for identifying high-risk patients.

## Introduction

Cardiovascular disease (CVD) remains the leading cause of death worldwide and a huge contributor to economic burden despite advancement of many excellent diagnoses and treatments. The prevalence of CVD continues to be high due to increasing life span in aging society, and the socio-economic costs are also continuously increasing. ^1, 2^ Many previous studies have identified traditional risk factors for CVD such as hypertension, dyslipidemia, diabetes, a family history of premature coronary artery disease (CAD), and smoking. ^3^ Recent studies have shown that socioeconomic status (SES) is also a factor related to CVD. ^4-6^ Although numerous studies on the relationship between SES and CVD have been conducted in the general population, ^4-6^ studies on the association between SES and cardiovascular outcomes in high-risk patients are scarce.

Cardiovascular events are more likely to occur in high-risk patients, so it is very important to identify their risk factors. Therefore, the purpose of this study was to investigate whether there is a difference in the presence and extent of coronary artery stenosis in invasive coronary angiography (ICA) according to SES in patients at high CVD risk. Since CAD severity is closely related to the occurrence of future cardiovascular events, we also intended to compare the prognosis according to SES.

## Methods

### Study subjects

Between May 2008 and May 2020, consecutive patients aged 20 to 90 years with suspected CAD who underwent ICA were retrospectively analyzed. There was no clinical limitation in enrollment except for patients diagnosed with cancer within the last five years. Finally, a total of 9,530 patients were analyzed in this study. This study was conducted in accordance with the Declaration of Helsinki, and the study protocol was approved by the Institutional Review Board (IRB) of Boramae Medical Center (Seoul, Korea) (IRB number, 10-2021-43). Obtaining written informed consent was waived by the IRB due to routine nature of information collected and retrospective nature of study design.

### Data collection

The patient’s height and body weight were measured at the time of admission. Body mass index (BMI) was calculated as weight in kilograms divided by the square of height in meters (kg/m^2^). History of cardiovascular risk factors, including, hypertension, diabetes mellitus, dyslipidemia, and current cigarette smoking status, was obtained. Previous clinical medical history, including myocardial infarction, coronary revascularization, heart failure, and stroke, was also assessed. Venous blood samples were obtained after overnight fasting for more than 12 hours and the following laboratory data were measured: hemoglobin, glycated hemoglobin (HbA1c), C-reactive protein, creatinine, total cholesterol, low-density lipoprotein (LDL) cholesterol, high-density lipoprotein (HDL) cholesterol, and triglyceride. Estimated glomerular filtration rate (eGFR) was calculated using the Modification of Diet in Renal Disease (MDRD) study equation. Transthoracic echocardiography was performed during hospitalization and left ventricular ejection fraction (LVEF) was measured with biplane method of disks (modified Simpson’s rule) according to the current practice guideline. ^7^ Information on current cardiovascular medications, including aspirin, clopidogrel, statins, angiotensin-converting enzyme inhibitors, angiotensin receptor blockers, beta-blockers, and calcium channel blockers, was also obtained.

### ICA and percutaneous coronary intervention

When performing ICA, the access site was determined at the operator’s discretion, regardless of either trans-radial or trans-femoral ICA. Obstructive CAD was defined as ≥50% stenosis in major coronary arteries or their branches with >2 mm diameter on ICA, and the extent of CAD based on the number of affected coronary arteries. Percutaneous coronary intervention (PCI) was performed according to current procedural guidelines. ^8-11^

### SES

The healthcare system in Korea is largely divided into national health insurance (NHI) and medical aid (MA), with private medical insurance serving as a supplementary function. NHI is a public health insurance program run by the Ministry of Health and Welfare that requires citizens with sufficient income to pay contributions to insure themselves and their dependents. The MA system is a public assistance system that the state guarantees for medical problems (disease, injury, childbirth, *etc*.) of low-income people who do not have the ability to sustain life or have difficulties in living. Medical aid beneficiaries (MABs) are households with less than 40% of the standard median income, households incapacitated to work, homeless people, facility recipients, victims of loss, and defectors from North Korea. ^12, 13^ Eligible amounts of household income for MAB are shown in **Supplementary Table S1**. In a previous study, compared to NHIB, MAB had lower income, lower education level, less participation in public pension, and lower private health insurance coverage. ^14^ In this study, the patients were divided into two groups according to the health insurance type: those with low SES who had the MA program (MAB group; n=1,436) and those with high SES who had the NHI program (NHIB group; n=8,094).

### Clinical events

The primary outcome was a major adverse cardiac and cerebral event (MACCE) which is a composite of cardiac death, non-fatal myocardial infarction, coronary revascularization, and non-fatal ischemic stroke during clinical follow-up. All deaths were considered to be related to cardiac causes unless a clear non-cardiac cause was identified. Myocardial infarction was defined as rise in cardiac troponin with at least one value above the 99th percentile upper reference limit and with at least one of the following: (1) symptoms of ischemia, (2) new or presumed new significant ST-segment–T wave changes or new left bundle branch block, (3) development of pathological Q waves in the electrocardiogram, or (4) imaging evidence of new loss of viable myocardium or new regional wall motion abnormality. ^15, 16^ Coronary revascularization was defined as revascularization with PCI or coronary artery bypass grafting. Ischemic stroke was defined as an episode of neurological dysfunction caused by focal cerebral infarction based on objective evidence of cerebral ischemic injury in a defined vascular distribution. ^17^

### Statistical analysis

The results are expressed as the mean ± standard deviation (SD) for continuous variables and as percentage for categorical variables. Between the two groups, continuous variables were compared using Student’s *t*-test and categorical variables were compared using Pearson’s chi-square test. The study endpoints were demonstrated using the Kaplan-Meier survival curve and compared using the log-rank test. Multivariable Cox proportional hazards models were used to determine whether medical insurance status was independently associated with clinical outcomes. Age, sex, BMI, hypertension, diabetes mellitus, smoking status, presence of obstructive CAD, LVEF, and the use of concomitant medications including antiplatelets, renin-angiotensin system blockers, beta-blockers, and statins, were included as covariates in the multivariable models. To determine whether the effect of medical insurance status differs depending on the presence or absence of CAD, clinical outcomes were also analyzed depending on the presence or absence of CAD. Global chi-square values were calculated to clarify incremental prognostic value of insurance type in combination with other risk factors for predicting future MACCE. All analyses were 2-tailed, and clinical significance was defined as *P*<0.05. Statistical analyses were performed with the statistical package SPSS version 23.0 (IBM Co., Armonk, NY, USA).

## Results

### Baseline characteristics and ICA-associated findings

The mean age of study subjects was 66.0±12.3 years, and 5,731 (60.2%) were male. The baseline clinical characteristics of the study subjects according to the health insurance type are shown in **Table 1**. Of the study patients, 1,436 (15.1%) were MABs. The MAB group was older (67.4±11.7 vs. 65.7±12.3 years; *P*<0.001) and had less male portion (57.2% vs. 60.8%; *P*=0.012) than the NHIB group. The MAB group had more cardiovascular risk factors, including hypertension, diabetes, and cigarette smoking, compared to the NHIB group (*P*<0.05 for each). Heart failure history was also more frequent in the MAB group than in the NIHB group. The diagnosis of acute myocardial infarction was more frequent in the NIHB group than in the MAB group. For major laboratory findings, hemoglobin and eGFR were lower, and glycated hemoglobin and C-reactive protein were higher in the MAB group than in the NHIB group. Mean LVEF was lower in the MAB group compared to the NHIB group. Among cardiovascular medications, statins were more frequently prescribed in the NHIB group, and angiotensin-converting enzyme inhibitors and beta-blockers were in the MAB group.

**Table 1.** Baseline characteristics of study subjects.

| **Characteristic** | **MAB**<br>**(n=1,436)** | **NHIB**<br>**(n=8,094)** | ***P* value** |
| --- | --- | --- | --- |
| Age, years | 67.4 ± 11.7 | 65.7 ± 12.3 | < 0.001 |
| Male | 822 (57.2) | 4,918 (60.8) | 0.012 |
| Body mass index, kg/m<sup>2</sup> | 24.2 ± 4.3 | 24.5 ± 3.6 | 0.045 |
| Cardiovascular risk factors |  |  |  |
| Hypertension | 937 (65.3) | 4,797 (59.3) | < 0.001 |
| Diabetes mellitus | 564 (39.3) | 2,540 (31.4) | < 0.001 |
| Dyslipidemia | 750 (52.2) | 4,478 (55.3) | 0.030 |
| Cigarette smoking | 444 (30.9) | 1,947 (24.1) | < 0.001 |
| Obesity (body mass index ≥ 25 kg/m<sup>2</sup>) | 535 (39.7) | 3,234 (42.2) | 0.087 |
| Medical history |  |  |  |
| Myocardial infarction | 81 (5.6) | 392 (4.8) | 0.200 |
| Coronary revascularization | 112 (7.8) | 519 (6.4) | 0.051 |
| Heart failure | 109 (7.6) | 448 (5.5) | 0.002 |
| Stroke | 133 (9.3) | 656 (8.1) | 0.143 |
| Clinical diagnosis |  |  |  |
| Acute myocardial infarction | 261 (18.1) | 1,760 (21.7) | 0.002 |
| Major laboratory findings |  |  |  |
| Hemoglobin, g/dL | 12.4 ± 2.0 | 13.0 ± 1.9 | < 0.001 |
| Glycated hemoglobin, % | 6.8 ± 1.7 | 6.5 ± 1.3 | < 0.001 |
| C-reactive protein, mg/dL | 1.9 ± 4.8 | 1.5 ± 4.0 | 0.015 |
| Estimated GFR, mL/min/1.73m<sup>2</sup> | 69.7 ± 33.8 | 76.5 ± 28.3 | < 0.001 |
| Total cholesterol, mg/dL | 159 ± 44 | 162 ± 43 | 0.012 |
| LDL cholesterol, mg/dL | 93.7 ± 37.4 | 94.6 ± 37.0 | 0.747 |
| HDL cholesterol, mg/dL | 43.4 ± 14.3 | 43.3 ± 12.4 | 0.850 |
| Triglyceride, mg/dL | 125 ± 79 | 123 ± 72 | 0.461 |
| Left ventricular ejection fraction, % | 58.2 ± 14.6 | 60.4 ± 13.3 | < 0.001 |
| Cardiovascular medications |  |  |  |
| Aspirin | 1,231 (85.7) | 6,840 (84.5) | 0.238 |
| Clopidogrel | 1,124 (78.3) | 6,204 (76.6) | 0.179 |
| Statins | 713 (49.7) | 4,250 (52.5) | 0.046 |
| Angiotensin-converting enzyme inhibitors | 163 (11.4) | 754 (9.3) | 0.016 |
| Angiotensin receptor blockers | 404 (28.1) | 2,114 (26.1) | 0.110 |
| Beta-blockers | 498 (34.7) | 2,521 (31.1) | 0.008 |
| Calcium channel blockers | 492 (34.3) | 2,546 (31.5) | 0.035 |

Numbers are expressed as mean ± standard deviation or n (%). MAB, medical aid beneficiaries; NHIB, National Health Insurance beneficiaries; GFR, glomerular filtration rate; HDL, high-density lipoprotein; LDL, low-density lipoprotein.

The ICA findings according to insurance status are shown in **Table 2**. Of the total study subjects, 6,100 (64.0%) had obstructive CAD. Overall, the prevalence and extent of obstructive CAD were not different between the two groups. The prevalence of left main disease and three-vessel disease was also similar between the two groups. The MAB group received PCI less frequently than the NHIB group (40.0% vs. 43.9%; *P*=0.006). The PCI results were similar between the two groups.

**Table 2.** ICA associated findings.

| **Characteristic** | **MAB**<br>**(n=1,436)** | **NHIB**<br>**(n=8,094)** | ***P* value** |
| --- | --- | --- | --- |
| Obstructive CAD | 902 (62.8) | 5,198 (64.2) | 0.306 |
| CAD extent |  |  | 0.045 |
| Insignificant | 534 (37.2) | 2,896 (35.8) |  |
| One-vessel disease | 253 (17.6) | 1,616 (20.0) |  |
| Two-vessel disease | 251 (17.5) | 1,530 (18.9) |  |
| Three-vessel disease | 398 (27.7) | 2,052 (25.4) |  |
| Left main disease | 94 (6.5) | 550 (6.8) | 0.729 |
| Three-vessel disease | 398 (27.7) | 2,052 (25.4) | 0.059 |
| Percutaneous coronary intervention | 574 (40.0) | 3,553 (43.9) | 0.006 |
| Procedure results |  |  | 0.551 |
| Successful | 535 (93.2) | 3,287 (92.5) |  |
| Suboptimal | 13 (2.3) | 110 (3.1) |  |
| Failed | 26 (4.5) | 156 (4.4) |  |

Numbers are expressed as n (%). ICA, invasive coronary angiography; MAB, medical aid beneficiaries; NHIB, National Health Insurance beneficiaries; CAD, coronary artery disease.

### Clinical events

The incidence of clinical events during the follow-up are shown in **Table 3**. During the median follow-up period of 3.5 years (interquartile range, 1.0 to 5.9 years), there were 1,598 (16.8%) MACCE. The incidence rate of MACCE was significantly higher in the MAB group than in the NHIB (20.2% vs. 16.2%, *P*<0.001), which was also confirmed in Kaplan-Meier survival curves (log-rank *P*<0.001) (**Figure 1**). In each component of MACCE, the incidence rates of cardiac death, myocardial infarction, ischemic stroke were not different between the two groups, but the rate of revascularization was significantly higher in the MAB group than in the NHIB group (10.8% vs. 6.9%, *P*<0.001). In the multivariable Cox regression analysis, medical insurance status was independently associated with the occurrence of MACCE during the follow-up period even after adjustment for multiple confounding factors (MAB vs. NHIB groups: hazard ratio [HR], 1.28; 95% confidence interval [CI], 1.07 to 1.54; *P*=0.006) (**Table 4**). Additionally, insurance status, age, diabetes mellitus, current smoking, reduced LVEF, and presence of obstructive CAD were associated with the occurrence of MACCE. To determine whether the effect of medical insurance status differs depending on the presence or absence of CAD, clinical events were also analyzed depending on the presence or absence of CAD. The incidence rate of MACCE was significantly higher in the MAB group than in the NHIB group, regardless of the presence or absence of CAD at baseline (log rank *P*=0.017 in patients without CAD, log rank *P*<0.001 in those with CAD) (**Figure 2**). In subgroups with (MAB vs. NHIB groups: HR, 1.24; 95% CI, 1.02 to 1.52; *P*=0.035) and without (MAB vs. NHIB groups: HR, 1.52; 95% CI, 1.01 to 2.29; *P*=0.047) CAD at baseline, medical insurance status was independently associated with the occurrence of MACCE during the follow-up period (**Table 5**). The global chi-square values of Cox regression analyses were compared to evaluate the incremental prognostic value of insurance status when it was added to age, sex, and traditional clinical factors including diabetes mellitus, hypertension, dyslipidemia, and smoking status. Addition of traditional clinical risk factors to age and sex had incremental prognostic value for predicting MACCE (global chi-square value, from 137.6 to 278.6; *P*<0.001). Addition of SES to combined use of age, sex, and traditional clinical factor results further increased the predictive power for MACCE (global chi-square value, from 278.6 to 285.5; *P*=0.008) (**Figure 3**).

**Table 3.** Clinical events.

| **Clinical event** | **MAB**<br>**(n=1,436)** | **NHIB**<br>**(n=8,094)** | ***P* value** |
| --- | --- | --- | --- |
| MACCE | 290 (20.2) | 1,308 (16.2) | < 0.001 |
| Cardiac death | 87 (6.1) | 478 (5.9) | 0.821 |
| Myocardial infarction | 24 (1.7) | 134 (1.7) | 0.966 |
| Coronary revascularization | 155 (10.8) | 560 (6.9) | < 0.001 |
| Ischemic stroke | 46 (3.2) | 203 (2.5) | 0.128 |

Numbers are expressed as n (%). MACCE, major adverse cardiac and cerebrovascular event;
MAB, medical aid beneficiaries; NHIB, National Health Insurance beneficiaries.

**Table 4.** Clinical factors associated with composite major cardiac and cerebrovascular events.

| Variable | HR | 95% CI | P value |
| --- | --- | --- | --- |
| MAB (vs. NHIB) | 1.28 | 1.07 to 1.54 | 0.006 |
| Age ≥ 65 years | 1.59 | 1.35 to 1.86 | < 0.001 |
| Female sex | 0.93 | 0.80 to 1.08 | 0.346 |
| BMI ≥ 25 kg/m<sup>2</sup> | 0.97 | 0.84 to 1.11 | 0.624 |
| Diabetes mellitus | 1.49 | 1.30 to 1.71 | < 0.001 |
| Hypertension | 1.07 | 0.91 to 1.25 | 0.400 |
| Current smoking | 1.26 | 1.06 to 1.49 | 0.008 |
| LVEF < 50% | 1.62 | 1.37 to 1.92 | < 0.001 |
| Obstructive CAD | 3.13 | 2.58 to 3.79 | < 0.001 |
| Antiplatelets | 0.92 | 0.72 to 1.19 | 0.543 |
| Statins | 0.90 | 0.78 to 1.04 | 0.162 |
| Beta-blockers | 1.03 | 0.90 to 1.18 | 0.686 |
| RAS blockers | 1.09 | 0.94 to 1.26 | 0.244 |

HR, hazard ratio; CI, confidence interval; MAB, medical aid beneficiaries; NHIB, National Health Insurance beneficiaries; BMI, body mass index; LVEF, left ventricular ejection fraction; CAD, coronary artery disease; RAS, renin angiotensin system.

**Table 5.** Clinical factors associated with composite major cardiac and cerebrovascular events according to presence of obstructive CAD.

| Variable | Without CAD (n=3,430) |  |  | With CAD (n=6,100) |  |  |
| --- | --- | --- | --- | --- | --- | --- |
|  | HR | 95% CI | P value | HR | 95% CI | P value |
| MAB (vs. NHIB) | 1.52 | 1.01 to 2.29 | 0.047 | 1.24 | 1.02 to 1.52 | 0.035 |
| Age ≥ 65 years | 2.22 | 1.52 to 3.26 | < 0.001 | 1.47 | 1.23 to 1.75 | < 0.001 |
| Female sex | 1.04 | 0.71 to 1.52 | 0.829 | 0.93 | 0.79 to 1.10 | 0.380 |
| BMI ≥ 25 kg/m<sup>2</sup> | 0.79 | 0.55 to 1.14 | 0.213 | 1.00 | 0.86 to 1.17 | 0.963 |
| Diabetes mellitus | 1.55 | 1.07 to 2.26 | 0.022 | 1.50 | 1.29 to 1.74 | < 0.001 |
| Hypertension | 1.16 | 0.79 to 1.72 | 0.449 | 1.05 | 0.89 to 1.25 | 0.550 |
| Current smoking | 1.57 | 1.00 to 2.47 | 0.050 | 1.21 | 1.01 to 1.46 | 0.037 |
| LVEF < 50% | 2.41 | 1.60 to 3.63 | < 0.001 | 1.55 | 1.29 to 1.86 | < 0.001 |
| Antiplatelets | 0.51 | 0.35 to 0.74 | < 0.001 | 1.45 | 1.00 to 2.11 | 0.052 |
| Statins | 0.70 | 0.49 to 1.01 | 0.053 | 0.93 | 0.80 to 1.10 | 0.398 |
| Beta-blockers | 0.78 | 0.53 to 1.14 | 0.199 | 1.06 | 0.91 to 1.23 | 0.447 |
| RAS blockers | 1.25 | 0.86 to 1.83 | 0.243 | 1.05 | 0.90 to 1.22 | 0.568 |

CAD, coronary artery disease; HR, hazard ratio; CI, confidence interval; MAB, medical aid beneficiaries; NHIB, National Health Insurance beneficiaries; BMI, body mass index; LVEF, left ventricular ejection fraction; RAS, renin angiotensin system.

**Figure 1.**
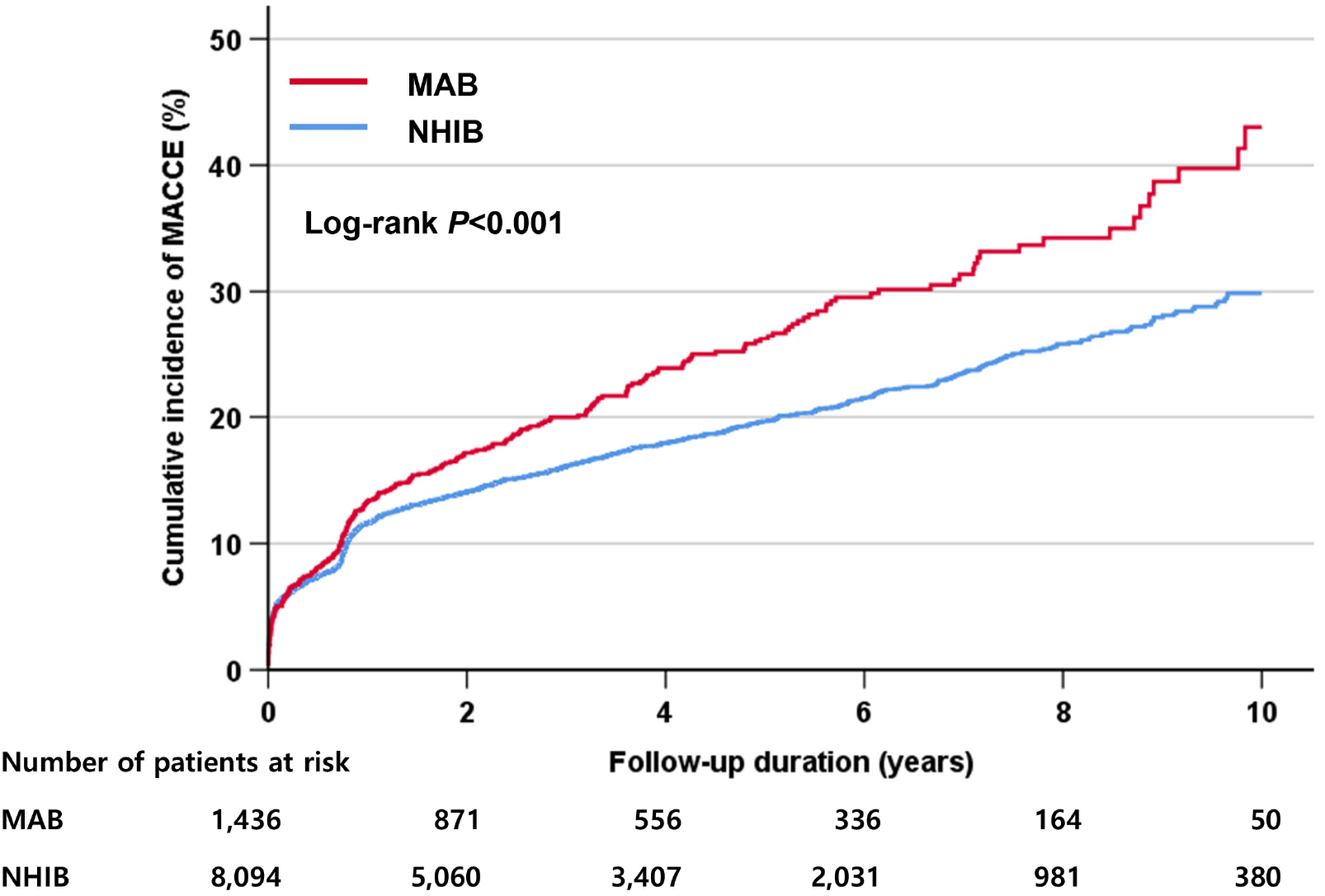
The incidence of MACCE according to the health insurance type. MACCE, major adverse cardiac and cerebral event; MAB, medical aid beneficiary; NHIB, national health insurance beneficiary.

**Figure 2.**
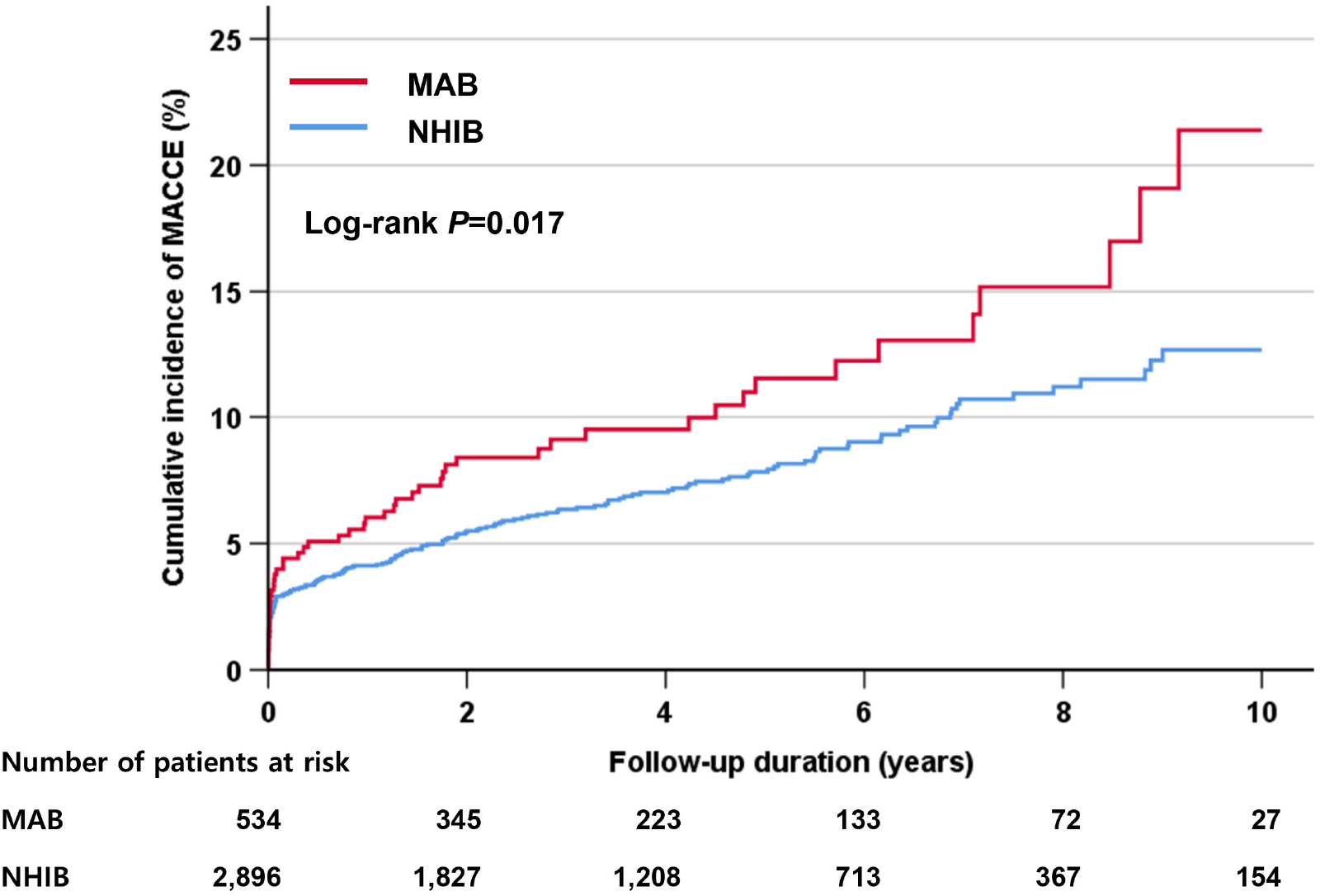
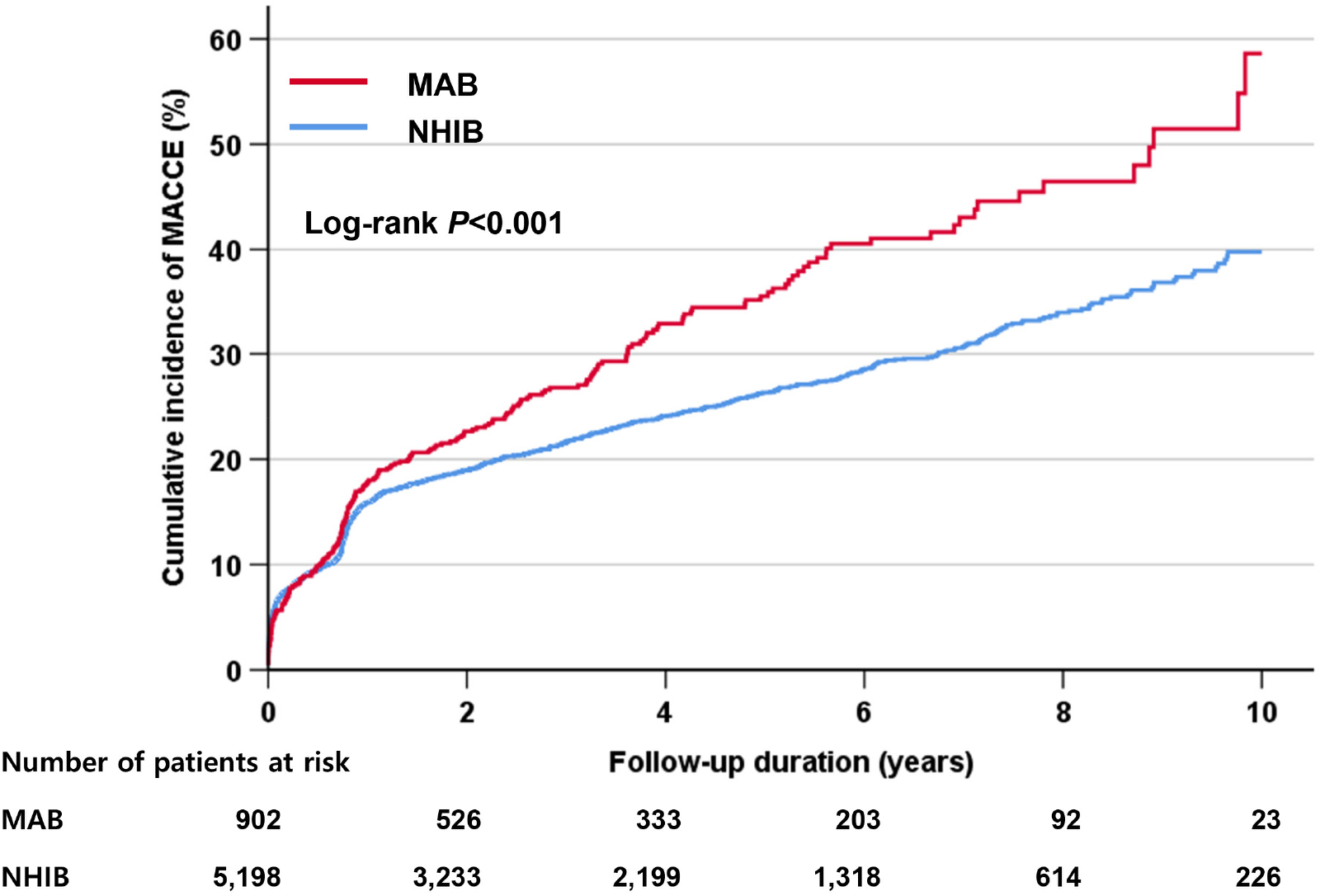
The incidence of MACCE in subjects with or without obstructive coronary artery disease according to the health insurance type. (a) Subjects without obstructive coronary artery disease, (b) subjects with obstructive coronary artery disease MACCE, major adverse cardiac and cerebral event; MAB, medical aid beneficiary; NHIB, national health insurance beneficiary.

**Figure 3.**
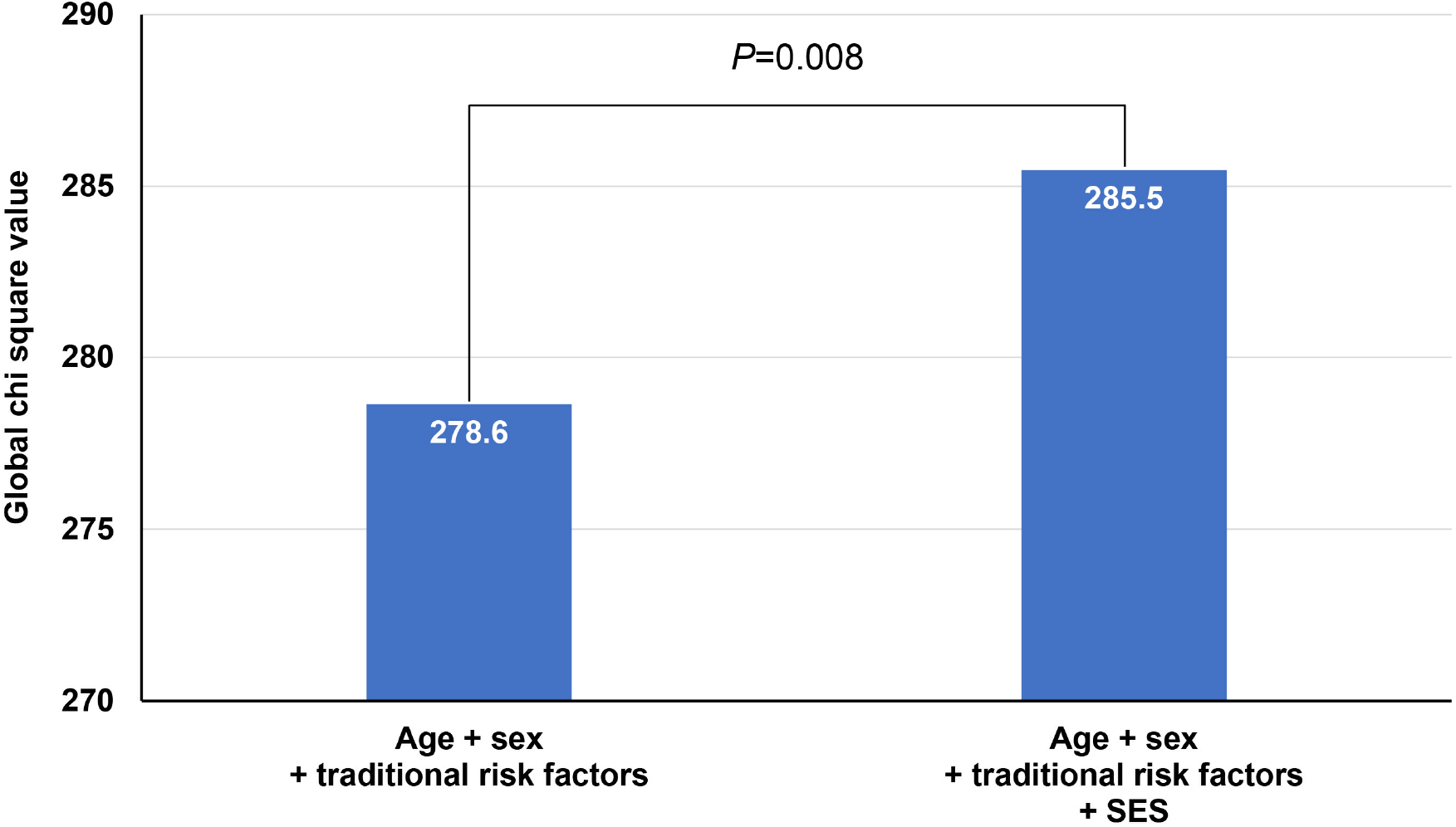
Prognostic value of clinical risk factors and health insurance type for MACCE. MACCE, major adverse cardiac and cerebral event.

## Discussion

The main finding of this study is that MABs had worse clinical outcomes than NHIBs in patients who underwent ICA. Although MABs were older and had more cardiovascular risk factors than NHIBs, the prevalence and extent of CAD were similar between the two groups. This study provided additional evidence for a relationship between low SES and increased CVD risk in a high-risk population.

Many previous studies have reported that low SES is associated with an increased risk of developing CVD and death. In previous study of Atherosclerosis Risk in Communities (ARIC) cohort consisting of the general population aged 45 to 64 years in four US communities and Finnish population-based cardiovascular risk factor monitoring cohorts (FINRISK), low income was significantly associated with increased risk of cardiac evets and sudden cardiac death. ^18^ In another study using ARIC data from more than 15,000 adults, low SES (low education and/or low income) was associated with older age, higher blood pressure, higher total cholesterol, more cigarette smoking, and more frequent use of antihypertensive medications. Thus, subjects with low SES had a higher Framingham risk score than those with high SES. In that study, low SES was associated with an increased coronary heart disease risk, independent of baseline Framingham risk score and time-dependent variables. ^19^ In an interesting computer simulation study of US adults (35 to 64 years) using data from the 2011- 2016 US National Health and Nutrition Examination Survey, both men and women in the low-SES group had double the rates of myocardial infarctions and cardiac deaths per 10,000 person-years compared to those in the high-SES group. The higher burden of traditional CAD risk factors in adults with low SES accounted for only 40% of these excess events. The remaining 60% of these events were attributed to other factors related to low SES. ^20^ In contrast to previous studies conducted in the general population, we studied the role of SES in high-risk patients with suspected CAD. In our study, similar to previous studies with the general population, low SES was associated with an increased risk of CVD in high-risk patients who underwent ICA for suspected CAD even after adjusting for baseline risk factors. Also, low SES was associated with an increased cardiovascular risk in both patients with and without obstructive CAD. The results of this study suggest that SES may be one of the significant risk factors for CVD, irrespective of baseline cardiovascular risk.

The exact mechanism by which SES acts as a risk factor for CVD is unclear, but there have been some studies that can infer the mechanism. According to a study with Korean National Health and Nutrition Examination Survey (KNHANES), low SES was associated with an increased risk of metabolic syndrome and its component in the Korean adult population. ^21^ Similarly, MABs had more cardiovascular risk factors, such as hypertension, diabetes, and smoking, in our study. In the Minnesota heart survey study, education level was inversely related to blood pressure, cigarette smoking, and BMI. In addition, a higher education level was associated with an increased time spent in leisure physical activities and health knowledge. ^22^ Lack of education can lead to risky decisions about health behaviors such as smoking and alcohol consumption. This suggest that SES is associated with modifiable behavioral risk factors. However, modifiable behavioral risk factors alone cannot explain the association between SES and CVD risk. The association between low SES and CVD risk is reinforced by an additional independent effect of SES. Another study suggested that low parental SES or low SES in childhood is associated with the onset of early CVD, independent of traditional risk factors. In a longitudinal cohort study of 1,337 medical students at Johns Hopkins School of Medicine, lower SES in childhood was associated with a higher risk of early coronary heart disease in adulthood among men with high adulthood SES. ^23^ As another factor, chronic psychosocial stress associated with low SES promotes atherosclerosis and CAD events. In the Whitehall II study with British civil servants, psychosocial work characteristics, and social support was associated with CAD incidence. ^24^ A systematic review of prospective cohort studies found that social isolation and lack of social support were associated with a two- to three-fold increased risk of cardiac mortality and morbidity. ^25^ In other study conducted in Denmark with more than 1,660,000 subjects, job strain was associated with a higher risk of developing CHD. ^26^ In addition to individual SES, socioeconomic characteristics of neighborhoods are associated with CVD risk factors, adverse events, and mortality. In the analysis using data from the Atherosclerosis Risk in Communities Study, living in disadvantaged areas is associated with an increased incidence of CHD after controlling for individual income, education, and occupation. ^27^ Collectively, low SES increases behavioral and psychosocial risks, and these disparities also occur as a result of poor health during childhood and early life, parental risk, and surrounding environments.

The poor prognosis of CVD in MABs is probably explained by the above reasons. This study did not enroll all MABs or NHIBs. However, in patients with suspected CAD who underwent ICA, baseline CAD status could be similar. Moreover, baseline underlying risk factors, such as hypertension, diabetes, and cigarette smoking, were more frequent in the MAB group than in the NHIB group. Although these risk factors were statistically adjusted, there may be a strong possibility that the control status of the patient’s risk factors and the duration of these risk factors would act as unadjusted factors. Besides this, interactions between behavioral and psychosocial risk, disparities during childhood, parental risk, and surrounding environments could lead worse CVD outcomes.

### Clinical implications

Subjects with low SES are more likely to develop CVD and to have a higher frequency of related risk factors, so CVD tests and treatments should be performed more aggressively. However, approaches only to clinical risk factors paradoxically increase the disparity as groups with higher SES are more likely to receive interventions in a healthcare setting. Therefore, appropriate risk stratification of low-SES patients with traditional risk factors for CVD is important for identifying high-risk patients. Although clear criteria on SES have not yet been established, SES should be considered a CVD risk factor. Furthermore, as modifiable behavioral factors and childhood disparities could influence future CVD outcomes, it is necessary to ensure proper health care and health education in childhood and to resolve social inequality caused by low SES.

### Study limitations

This study has several limitations. First, as this study is a single-center study in Korea, it is difficult to generalize our results to other races and populations. Secondly, as this study is of retrospective design, there is a possibility that clinical event information may be inaccurate. Thirdly, even within the same MAB or NHIB groups, educational level, income, and social position can vary widely among individuals. Finally, changes in cardiovascular risk factors and medications were not investigated during the follow-up period.

### Conclusions

Although CAD prevalence was similar, MABs showed an increased risk of composite cardiovascular events than NHIBs in Korean adults undergoing ICA. This provides additional evidence for the association between low SES and an increased risk of CVD even in populations at high risk. Cardiologists need an elaborate long-term strategy to reduce cardiovascular risk factors in low-SES patients.

## Source of Funding

None.

## Disclosures

The authors declare that the research was conducted in the absence of any commercial or financial relationships that could be construed as a potential conflict of interest.

## Data Availability

The datasets generated during and/or analysed during the current study are available from the corresponding author on reasonable request.

## Notes

### Competing Interest Statement

The authors have declared no competing interest.

### Author Declarations

The Institutional Review Board (IRB) of Boramae Medical Center (Seoul, Korea) (IRB number, 10-2021-43).

